# A framework for classifying disease trends applied to influenza-associated hospital admissions in the United States

**DOI:** 10.1101/2025.04.11.25324947

**Authors:** Sarabeth M. Mathis, Matthew Biggerstaff, Alicia Budd, Alissa O’Halloran, Catherine Bozio, Rebecca K. Borchering

## Abstract

We built a framework for categorizing week-to-week increases or decreases in seasonal influenza hospitalizations aiding data interpretation in the context of past seasons. Using Influenza Hospitalization Surveillance Network (FluSurv-NET) data, we established thresholds for weekly hospitalization rate differences and applied them to the 2022/23 and 2023/24 influenza seasons from the National Healthcare Safety Network. While the number of weeks categorized as stable was consistent across seasons, more large increases and decreases were observed in 2022/23. This metric captures hospitalization rate changes and contextualizes the current season relative to past influenza trends, facilitating trend assessment.

## Introduction

Seasonal influenza causes substantial morbidity and mortality.[1]. In the United States, seasonal influenza laboratory test results, hospitalizations, and adult deaths are not nationally notifiable. Therefore, a variety of surveillance systems are used to monitor influenza activity including the Influenza Hospitalization Surveillance Network (FluSurv-NET) and the National Healthcare Safety Network (NHSN) [2]. These surveillance systems can inform where influenza activity is happening and how many people have tested positive for influenza or had influenza-associated hospitalizations. However, these indicators often require broader context to understand the overall population-level impact of influenza. The Moving Epidemic Method (MEM) and the Average Curve Method (ACM), among other approaches, were developed to help provide this context[3]. These tools have been used across the globe to set thresholds for determining the severity of influenza and other respiratory viruses [4-6]. CDC uses the MEM approach to help classify the severity of seasonal influenza epidemics[4].

In addition to the overall impact of seasonal influenza, a key public health question is also whether influenza activity is stable over time or changing (i.e., trends in influenza activity). Quantitatively evaluating recent weeks’ trends during the current season may aid in early detection of atypical increases or unusual patterns in influenza activity. Quantitatively characterized weekly disease trend levels relative to what has been observed during previous seasons can provide a strong foundation for situational awareness and help inform assessments of current activity and where unusual increases in influenza activity are occurring. Here we describe efforts to develop a method to classify trends in influenza activity based on historic data and describe influenza hospital admissions trends across two seasons in the United States.

## Methods

We analyzed hospital admission data from two surveillance systems, FluSurv-NET and NHSN. FluSurv-NET is a population-based surveillance system that collects data on laboratory-confirmed influenza associated hospital admissions among children and adults [7]. Active surveillance is conducted from October 1 through April 30 every year but was extended to June 11, 2022, during the 2021/22 season due to late influenza season activity. Detailed FluSurv-NET methods are available elsewhere[8, 9]. Weekly rates per 100,000 population are available from participating sites as well as an overall network rate[2]. As of the 2022/23 season, FluSurv-NET includes more than 90 counties in 14 states and an estimated nine percent of the US population.

The NHSN hospital admission dataset refers to the COVID-19 Reported Patient impact and Hospital Capacity by State Timeseries (originally called HHS-Protect)[10]. During the COVID-19 pandemic, HHS mandated that hospitals report daily influenza hospital admission counts in each state and Washington DC beginning February 2, 2022. The original mandate remained in effect through April 30, 2024[11]. For this analysis, daily influenza hospital admission counts were aggregated by state and epidemiological week and converted to per capita rates using 2021 and 2022 US Census population estimates[12, 13].

From FluSurv-NET, starting with the 2010/11 season through 2022/23, excluding 2020/21, and from NHSN from 2021/22 through 2023/24, we calculated week-to-week differences in influenza hospital admission rates for each season. For both datasets, we began including rate differences in our observed difference distributions once there had been at least three consecutive weeks of non-zero rates at the start of each season, to capture changes in influenza activity during transmission periods.

From the distribution of rate differences, we established five categories of change: stable, increase, large increase, decrease, large decrease. Rounded FluSurv-NET rate differences were used to specify the category thresholds. Differences that fell roughly between the 25^th^ percentile and 75^th^ percentile of the distribution were classified as stable, differences from 75^th^ and 95^th^ percentile, were classified as increase, 5^th^ to 25^th^ percentile were classified as decrease, and differences lower than or equal to the 5^th^ percentile or greater than or equal to the 95^th^ percentiles were classified as large decrease or large increase respectively. For each jurisdiction, weeks with an absolute change of less than 10 admissions were classified as stable to limit the impact of reporting delays and noise (most NHSN influenza hospital admissions counts were revised by less than 10 admissions per week after initial publication [14]). We applied the rate thresholds, developed using FluSurv-NET, to NHSN hospital admission data for each jurisdiction to characterize the changes observed throughout the most recent influenza seasons.

## Results

### Historic data analysis and threshold determination

The median rate difference between consecutive weeks of historic FluSurv-NET and NHSN data was 0 influenza-associated hospitalizations per 100,000 individuals. The distribution of rate differences for both datasets was roughly symmetric around the median (Supplement Table 1 and Supplemental Figure 1). Supplement Table 1 shows mean, standard deviation, median, range, and percentiles for the rate differences from FluSurv-NET and NHSN.

Based on the criteria described in the methods, stable weeks were categorized as a change in weekly rate greater than -0.3 influenza-associated hospitalizations/100,000 but less than 0.3, or weeks with an absolute difference of fewer than 10 influenza hospital admissions (Table 1).

**Table 1.**
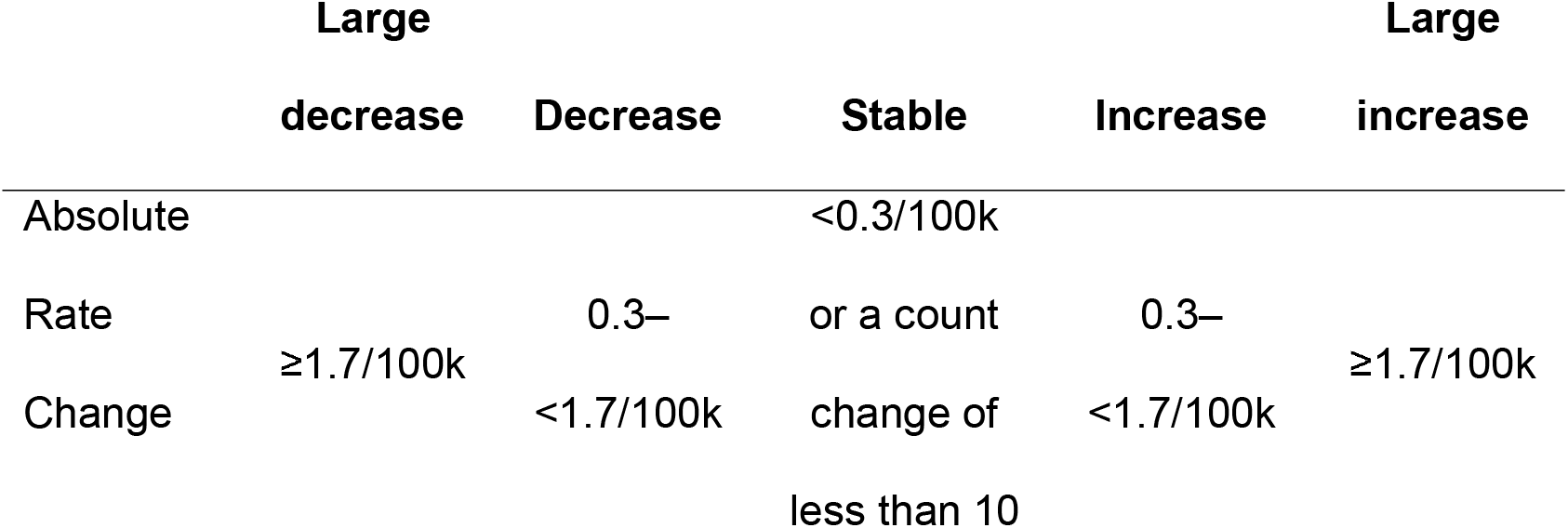
Absolute influenza rate and count thresholds for category specifications based on updated percentiles. Negative changes that did not qualify as stable are classifed as decreases and large decreases and positive changes that did not qualify as stable are classified as increases or large increases.

Increases were categorized as a positive change in weekly rates from 0.3 to less than 1.7, and large increases as a rate change of 1.7/100,000 influenza-associated hospitalizations or greater. Decreases and large decreases were categorized for negative rate changes using the same increase and large increase thresholds described above.

### Framework applied to 2022/23 and 2023/24 influenza seasons

Figure 1 shows national (panel a) and US jurisdictions (panel b) across the 2022/23 and 2023/24 seasons with weeks colored by week-to-week trend category. The 22/23 influenza season was characterized by a sharp peak of influenza-associated hospitalizations in early December and a rapid decrease in hospital admissions with low activity throughout the rest of the season in most of the US. In the 2022/23 season, 7.4% and 7.2% of weeks were classified as large decreases and large increases each, 12.6% and 12.5% of weeks were classified as decreases and increases, respectively. The week with the most observed increases and large increases was 11/26/2022 when 14 states observed increases and 24 states observed large increases. The week of January 14, 2023, had the most observed decreases or large decreases when 19 states observed decreases and 32 states observed large decreases. From 2/11/2023 through 5/27/2023, more than 40 states observed stable trends each week.

**Figure 1.**
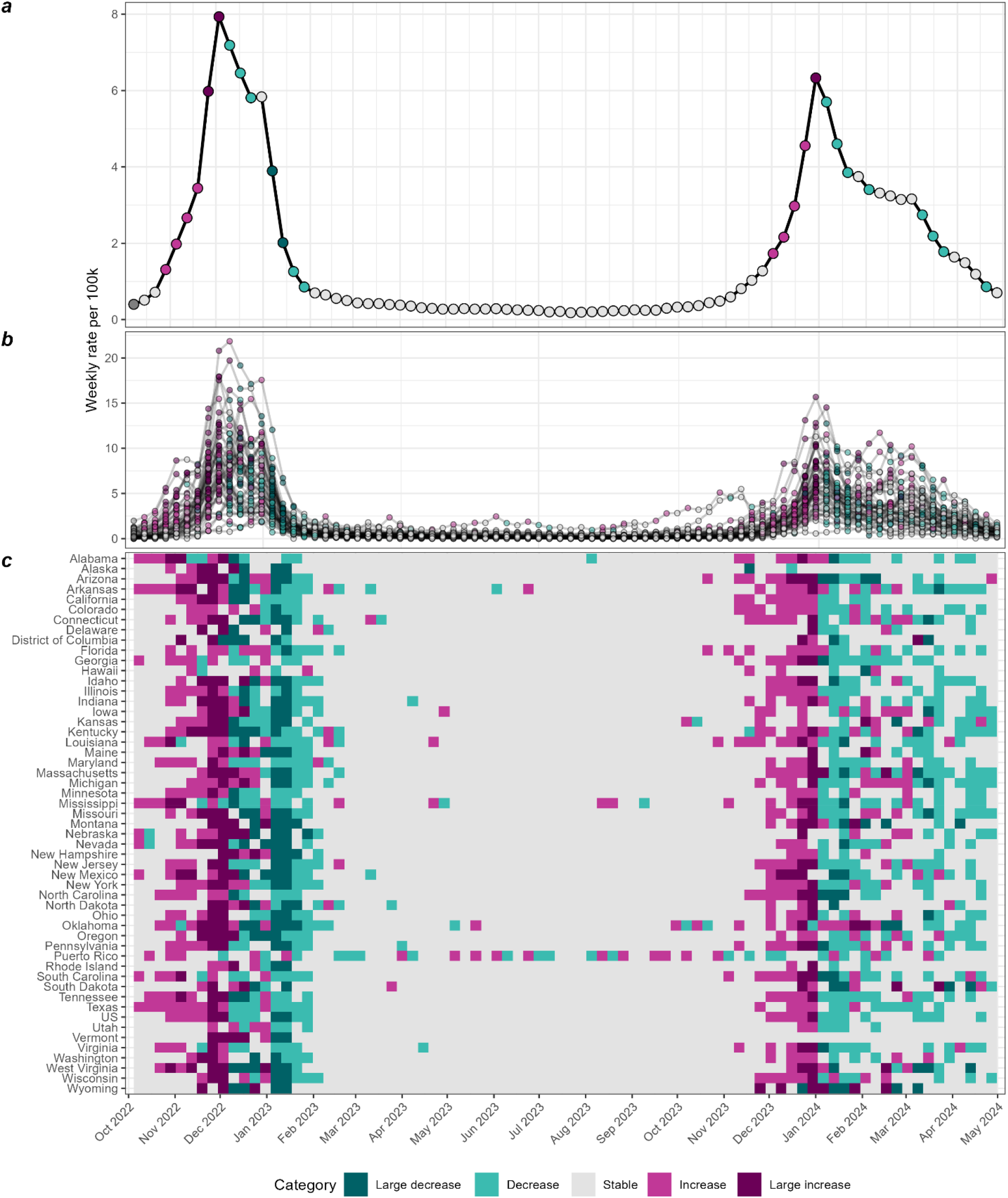
Week-to-week influenza hospitalization rate change trends for NHSN 2022/23 and 2023/24 seasons. Panels a and b show weekly rates per 100k colored by trend category for the US (panel a) and each state, Puerto Rico and Washington DC (panel b). Points are colored by trend category based on the rate-difference from the previous week. Panel c shows the week-to-week trend categories by jurisdiction. The dark gray at the beginning of each panel indicates that not enough data were available to categorize that week.

A similar but slightly less sharp peak occurred in late December of the 2023/24 season with a much slower overall decline in activity and a second peak in some jurisdictions later in the season. The 23/24 season had 3.3% and 4.2% of weeks classified as large decreases and large increases, respectively, 19.4% as decreases and 16.1% of weeks classified as increases. Two weeks shared the most observed increases and large increases. The week of 12/23/2023 observed 25 increases and 18 large increases and the week of 12/30/2023 observed 14 increases and 29 large increases. The week with the most decreases and large decreases was 1/13/24 when 30 states observed decreases and 11 observed large decreases. The count of stable jurisdictions fluctuated between 18 and 23 through 4/27/24.

When applied to week 40 through week 17 of the 2022/23 and 2023/24 seasons, 60% and 57% of weeks across all jurisdictions were considered stable for each season, respectively.

Supplemental Tables 1 and 2 detail the distributions of categorized weeks by jurisdiction for week-to-week rate differences for the 22/23 and 23/24 seasons, respectively.

## Discussion

In this analysis, we used historic FluSurv-NET weekly data to determine thresholds to categorize changes in NHSN influenza hospital admission trends due to seasonal influenza. We aimed to create robust classifications that enhanced our understanding of influenza activity over multiple seasons in the United States and applied our rate change classifications to two seasons of NHSN data to understand how this metric would perform during seasonal influenza epidemics. These categories proved to be useful for characterizing week-to-week trends in influenza activity. In addition, this method complemented other existing methods to understand the impact of influenza, such as MEM and ACM [10, 11], which require more extensive epidemiological and statistical modeling knowledge and do not determine if changes in metrics differ from expectations based on past seasons.

The metrics developed in this analysis can be deployed in real-time to enhance the interpretation of surveillance data, detect changes in influenza activity, and help guide efforts to understand potential contributing factors. Selecting larger percentiles to base the change metrics on or using other surveillance indicators (e.g., influenza test positivity) to establish trend classifications could enhance the ability of this approach to detect more atypical changes in activity. Beyond interpreting surveillance data, the trend classifications can also be used to interpret forecasts or to directly forecast probability of the trend categories occurring over the coming weeks[15]. This application provides a new way to communicate forecasts that could enhance their interpretability and utility within public health contexts.

Robust and sustained surveillance systems are needed to create trend classifications that are not subject to effects of changes in reporting while also capturing the range of activity that can be observed over multiple influenza seasons with varying intensity. Being able to use several seasons of FluSurv-NET data, alongside the more recently available NHSN data, helped make the definitions more robust than just using two and a half seasons of available NHSN data or data from the 14 FluSurv-NET jurisdictions alone to set the thresholds.

In summary, our analysis highlights the capability of utilizing historical data from various sources to establish thresholds for classifying changes in seasonal influenza activity. By developing a flexible framework that integrates multiple data sources while remaining accessible to public health practitioners, we aim to enhance real-time surveillance efforts. This approach not only improves trend detection but can aid in forecasting future disease activity, and ultimately support more effective public health responses during influenza seasons. For example, accurate forecasts of increases could help hospitals prepare for larger influxes of patients with influenza. While our analysis was focused on changes in influenza hospital admission rates, we believe this framework could be used with other surveillance systems or metrics like influenza emergency department visits or percent positivity. It could also be adapted to other diseases where appropriately long and robust time-series data are available with activity levels to characterize a variety of increase and decreasing periods of disease activity. Additionally, we plan to refine our classification metrics through ongoing evaluation to ensure that they remain relevant amid evolving epidemiological landscapes.

## Data Availability

All data produced in the present study are available upon reasonable request to the authors. All data used in the present study are available online at cdc.gov and healthdata.gov

https://gis.cdc.gov/GRASP/Fluview/FluHospRates.html

https://healthdata.gov/Hospital/COVID-19-Reported-Patient-Impact-and-Hospital-Capa/g62h-syeh/about_data

## Disclaimer

The findings and conclusions in this report are those of the authors and do not necessarily represent the official position of the Centers for Disease Control and Prevention.

## Author contributions

S.M.M., R.K.B., and M.B. conceptualized the analysis. S.M.M. and R.K.B. developed the methodology. A.O’H. and C.B. curated FluSurv-NET data. S.M.M. performed data analyses and A.B., M.B., and R.K.B assisted with interpretation of results. S.M.M. wrote initial draft. S.M.M., M.B., A.B., A.O’H., C.B., and R.K.B. participated in reviewing and editing the manuscript. All authors reviewed and approved the final version before submission.

