## Supplemental Tables and Figure for "A framework for classifying disease trends applied to influenza-associated hospital admissions in the United States"

**Supplement Figure 1. Week-to-week influenza hospitalization rate change histogram by dataset, colored by trend category.** FluSurv-NET seasons include 2010/11 – 2022/23, except 2020/21, NHSN seasons include 2021/22 – 2022/23. The dashed lines are showing the percentiles, in order from left to right are $5^{th}$, $12.5^{th}$, $25^{th}$, $37.5^{th}$, $62.5^{th}$, $75^{th}$, $87.5^{th}$, and $95^{th}$. Supplmental Table 1 shows the values associated with each percentile from each dataset. The graph/histogram bars are colored by the eventually determined category showing how much of the histogram would be considered each category. Not shown are the count thresholds. Weeks with a difference of fewer than 10 hospital admissions from the previous week would be considered stable, otherwise they would be categorized according to the rate difference thresholds.

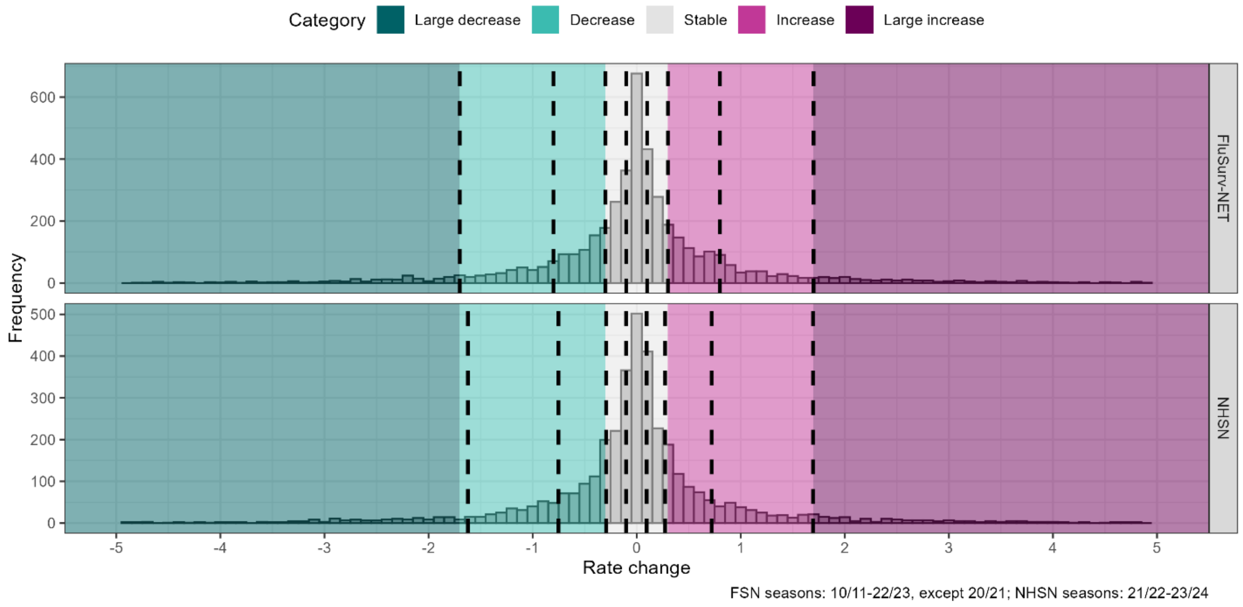

**Supplement Table 1. Influenza hospitalization rate difference statistics by dataset and horizon.** This table shows the percentiles of each rate difference distribution by dataset and horizon. The $5^{th}$ and $95^{th}$ percentiles contain the lower and upper bounds or 90% of the rate differences per dataset and horizon, the $12.5^{th}$ and $87.5^{th}$ percentiles contain the lower and upper bounds for 75% of the data, the $25^{th}$ and $75^{th}$ percentiles contain the lower and upper bounds for 50% of the data, and the $37.5^{th}$ and $62.5^{th}$ bound 25% of the data.

| | | | | **Percentiles** | | | | | | | |
|---|---|---|---|---|---|---|---|---|---|---|---|
| **Dataset** | **Week difference** | **Mean (SD)** | **Median (range)** | **5** | **12.5** | **25** | **37.5** | **62.5** | **75** | **87.5** | **95** |
| FluSurv-NET | 1 | 0 (1.2) | 0 (-9.9, 16.1) | -1.7 | -0.8 | -0.3 | -0.1 | 0.1 | 0.3 | 0.8 | 1.7 |
| NHSN | 1 | 0 (1.3) | 0 (-6.2, 10.1) | -1.6 | -0.8 | -0.3 | -0.1 | 0.1 | 0.3 | 0.7 | 1.7 |

**Supplemental Table 2 Distribution of influenza trend categories by jurisdiction for the 2022/23 season.** Weeks 40 through 17 from NHSN were included.

| **Jurisdiction** | **Large decrease** N (%) | **Decrease** N (%) | **Stable** N (%) | **Increase** N (%) | **Large increase** N (%) |
|---|---|---|---|---|---|
| | 117 (7.4%) | 200 (12.6%) | 960 (60.4%) | 199 (12.5%) | 114 (7.2%) |
| US | 2 (6.7%) | 5 (16.7%) | 17 (56.7%) | 4 (13.3%) | 2 (6.7%) |
| Alabama | 1 (3.3%) | 5 (16.7%) | 17 (56.7%) | 4 (13.3%) | 3 (10%) |
| Alaska | 3 (10%) | 0 (0%) | 22 (73.3%) | 2 (6.7%) | 3 (10%) |
| Arizona | 3 (10%) | 3 (10%) | 17 (56.7%) | 4 (13.3%) | 3 (10%) |
| Arkansas | 3 (10%) | 6 (20%) | 11 (36.7%) | 7 (23.3%) | 3 (10%) |
| California | 2 (6.7%) | 4 (13.3%) | 19 (63.3%) | 3 (10%) | 2 (6.7%) |
| Colorado | 1 (3.3%) | 3 (10%) | 21 (70%) | 5 (16.7%) | 0 (0%) |
| Connecticut | 4 (13.3%) | 4 (13.3%) | 16 (53.3%) | 4 (13.3%) | 2 (6.7%) |
| Delaware | 2 (6.7%) | 1 (3.3%) | 23 (76.7%) | 2 (6.7%) | 2 (6.7%) |

| Jurisdiction | Large decrease N (%) | Decrease N (%) | Stable N (%) | Increase N (%) | Large increase N (%) |
|---|---|---|---|---|---|
| District of Columbia | 4 (13.3%) | 1 (3.3%) | 23 (76.7%) | 0 (0%) | 2 (6.7%) |
| Florida | 1 (3.3%) | 6 (20%) | 16 (53.3%) | 7 (23.3%) | 0 (0%) |
| Georgia | 0 (0%) | 5 (16.7%) | 19 (63.3%) | 6 (20%) | 0 (0%) |
| Hawaii | 0 (0%) | 2 (6.7%) | 25 (83.3%) | 3 (10%) | 0 (0%) |
| Idaho | 3 (10%) | 3 (10%) | 18 (60%) | 2 (6.7%) | 4 (13.3%) |
| Illinois | 2 (6.7%) | 4 (13.3%) | 18 (60%) | 5 (16.7%) | 1 (3.3%) |
| Indiana | 3 (10%) | 5 (16.7%) | 16 (53.3%) | 3 (10%) | 3 (10%) |
| Iowa | 2 (6.7%) | 4 (13.3%) | 18 (60%) | 3 (10%) | 3 (10%) |
| Kansas | 3 (10%) | 4 (13.3%) | 18 (60%) | 3 (10%) | 2 (6.7%) |
| Kentucky | 4 (13.3%) | 5 (16.7%) | 14 (46.7%) | 3 (10%) | 4 (13.3%) |
| Louisiana | 1 (3.3%) | 7 (23.3%) | 14 (46.7%) | 7 (23.3%) | 1 (3.3%) |
| Maine | 3 (10%) | 2 (6.7%) | 20 (66.7%) | 3 (10%) | 2 (6.7%) |
| Maryland | 0 (0%) | 8 (26.7%) | 15 (50%) | 6 (20%) | 1 (3.3%) |
| Massachusetts | 2 (6.7%) | 4 (13.3%) | 18 (60%) | 3 (10%) | 3 (10%) |
| Michigan | 2 (6.7%) | 3 (10%) | 18 (60%) | 5 (16.7%) | 2 (6.7%) |
| Minnesota | 1 (3.3%) | 5 (16.7%) | 18 (60%) | 5 (16.7%) | 1 (3.3%) |
| Mississippi | 1 (3.3%) | 8 (26.7%) | 12 (40%) | 7 (23.3%) | 2 (6.7%) |
| Missouri | 4 (13.3%) | 2 (6.7%) | 18 (60%) | 3 (10%) | 3 (10%) |
| Montana | 4 (13.3%) | 0 (0%) | 22 (73.3%) | 0 (0%) | 4 (13.3%) |
| Nebraska | 4 (13.3%) | 2 (6.7%) | 17 (56.7%) | 4 (13.3%) | 3 (10%) |
| Nevada | 4 (13.3%) | 3 (10%) | 17 (56.7%) | 3 (10%) | 3 (10%) |
| New Hampshire | 3 (10%) | 1 (3.3%) | 21 (70%) | 2 (6.7%) | 3 (10%) |
| New Jersey | 2 (6.7%) | 6 (20%) | 16 (53.3%) | 4 (13.3%) | 2 (6.7%) |

| Jurisdiction | Large decrease<br>N (%) | Decrease<br>N (%) | Stable<br>N (%) | Increase<br>N (%) | Large increase<br>N (%) |
|---|---|---|---|---|---|
| New Mexico | 4 (13.3%) | 3 (10%) | 16 (53.3%) | 3 (10%) | 4 (13.3%) |
| New York | 2 (6.7%) | 5 (16.7%) | 16 (53.3%) | 5 (16.7%) | 2 (6.7%) |
| North Carolina | 1 (3.3%) | 4 (13.3%) | 19 (63.3%) | 6 (20%) | 0 (0%) |
| North Dakota | 3 (10%) | 4 (13.3%) | 17 (56.7%) | 3 (10%) | 3 (10%) |
| Ohio | 2 (6.7%) | 4 (13.3%) | 19 (63.3%) | 3 (10%) | 2 (6.7%) |
| Oklahoma | 5 (16.7%) | 4 (13.3%) | 13 (43.3%) | 5 (16.7%) | 3 (10%) |
| Oregon | 3 (10%) | 4 (13.3%) | 18 (60%) | 1 (3.3%) | 4 (13.3%) |
| Pennsylvania | 2 (6.7%) | 5 (16.7%) | 17 (56.7%) | 4 (13.3%) | 2 (6.7%) |
| Puerto Rico | 0 (0%) | 7 (23.3%) | 17 (56.7%) | 6 (20%) | 0 (0%) |
| Rhode Island | 2 (6.7%) | 1 (3.3%) | 23 (76.7%) | 3 (10%) | 1 (3.3%) |
| South Carolina | 0 (0%) | 7 (23.3%) | 17 (56.7%) | 5 (16.7%) | 1 (3.3%) |
| South Dakota | 2 (6.7%) | 1 (3.3%) | 22 (73.3%) | 3 (10%) | 2 (6.7%) |
| Tennessee | 2 (6.7%) | 4 (13.3%) | 16 (53.3%) | 5 (16.7%) | 3 (10%) |
| Texas | 0 (0%) | 7 (23.3%) | 13 (43.3%) | 9 (30%) | 1 (3.3%) |
| Utah | 0 (0%) | 4 (13.3%) | 22 (73.3%) | 4 (13.3%) | 0 (0%) |
| Vermont | 2 (6.7%) | 0 (0%) | 23 (76.7%) | 1 (3.3%) | 4 (13.3%) |
| Virginia | 0 (0%) | 7 (23.3%) | 18 (60%) | 4 (13.3%) | 1 (3.3%) |
| Washington | 2 (6.7%) | 3 (10%) | 21 (70%) | 2 (6.7%) | 2 (6.7%) |
| West Virginia | 5 (16.7%) | 1 (3.3%) | 17 (56.7%) | 2 (6.7%) | 5 (16.7%) |
| Wisconsin | 2 (6.7%) | 4 (13.3%) | 19 (63.3%) | 3 (10%) | 2 (6.7%) |
| Wyoming | 4 (13.3%) | 0 (0%) | 23 (76.7%) | 0 (0%) | 3 (10%) |

**Supplemental Table 3 Distribution of influenza hospitalization trend categories by jurisdiction for the 2023/24 season.** Weeks 40 through 17 from NHSN were included.

| Jurisdiction | Large decrease N (%) | Decrease N (%) | Stable N (%) | Increase N (%) | Large increase N (%) |
|---|---|---|---|---|---|
| | 53 (3.3%) | 309 (19.4%) | 906 (57.0%) | 256 (16.1%) | 66 (4.2%) |
| US | 0 (0 %) | 8 (26.7 %) | 17 (56.7 %) | 4 (13.3 %) | 1 (3.3 %) |
| Alabama | 1 (3.3 %) | 7 (23.3 %) | 15 (50 %) | 6 (20 %) | 1 (3.3 %) |
| Alaska | 1 (3.3 %) | 1 (3.3 %) | 28 (93.3 %) | 0 (0 %) | 0 (0 %) |
| Arizona | 4 (13.3 %) | 4 (13.3 %) | 13 (43.3 %) | 6 (20 %) | 3 (10 %) |
| Arkansas | 1 (3.3 %) | 9 (30 %) | 11 (36.7 %) | 8 (26.7 %) | 1 (3.3 %) |
| California | 0 (0 %) | 4 (13.3 %) | 19 (63.3 %) | 7 (23.3 %) | 0 (0 %) |
| Colorado | 0 (0 %) | 9 (30 %) | 13 (43.3 %) | 8 (26.7 %) | 0 (0 %) |
| Connecticut | 1 (3.3 %) | 7 (23.3 %) | 12 (40 %) | 9 (30 %) | 1 (3.3 %) |
| Delaware | 0 (0 %) | 4 (13.3 %) | 24 (80 %) | 1 (3.3 %) | 1 (3.3 %) |
| District of Columbia | 2 (6.7 %) | 1 (3.3 %) | 24 (80 %) | 1 (3.3 %) | 2 (6.7 %) |
| Florida | 0 (0 %) | 8 (26.7 %) | 14 (46.7 %) | 8 (26.7 %) | 0 (0 %) |
| Georgia | 1 (3.3 %) | 9 (30 %) | 14 (46.7 %) | 4 (13.3 %) | 2 (6.7 %) |
| Hawaii | 1 (3.3 %) | 2 (6.7 %) | 24 (80 %) | 3 (10 %) | 0 (0 %) |
| Idaho | 1 (3.3 %) | 8 (26.7 %) | 15 (50 %) | 4 (13.3 %) | 2 (6.7 %) |
| Illinois | 0 (0 %) | 7 (23.3 %) | 18 (60 %) | 5 (16.7 %) | 0 (0 %) |
| Indiana | 0 (0 %) | 8 (26.7 %) | 15 (50 %) | 6 (20 %) | 1 (3.3 %) |
| Iowa | 1 (3.3 %) | 6 (20 %) | 15 (50 %) | 8 (26.7 %) | 0 (0 %) |
| Kansas | 1 (3.3 %) | 6 (20 %) | 16 (53.3 %) | 6 (20 %) | 1 (3.3 %) |
| Kentucky | 1 (3.3 %) | 9 (30 %) | 11 (36.7 %) | 8 (26.7 %) | 1 (3.3 %) |
| Louisiana | 1 (3.3 %) | 7 (23.3 %) | 13 (43.3 %) | 8 (26.7 %) | 1 (3.3 %) |

| Jurisdiction | Large decrease N (%) | Decrease N (%) | Stable N (%) | Increase N (%) | Large increase N (%) |
|---|---|---|---|---|---|
| Maine | 0 (0 %) | 4 (13.3 %) | 23 (76.7 %) | 1 (3.3 %) | 2 (6.7 %) |
| Maryland | 0 (0 %) | 7 (23.3 %) | 18 (60 %) | 4 (13.3 %) | 1 (3.3 %) |
| Massachusetts | 2 (6.7 %) | 10 (33.3 %) | 9 (30 %) | 6 (20 %) | 3 (10 %) |
| Michigan | 1 (3.3 %) | 9 (30 %) | 11 (36.7 %) | 8 (26.7 %) | 1 (3.3 %) |
| Minnesota | 0 (0 %) | 5 (16.7 %) | 20 (66.7 %) | 5 (16.7 %) | 0 (0 %) |
| Mississippi | 1 (3.3 %) | 9 (30 %) | 14 (46.7 %) | 4 (13.3 %) | 2 (6.7 %) |
| Missouri | 1 (3.3 %) | 9 (30 %) | 13 (43.3 %) | 6 (20 %) | 1 (3.3 %) |
| Montana | 4 (13.3 %) | 2 (6.7 %) | 18 (60 %) | 3 (10 %) | 3 (10 %) |
| Nebraska | 0 (0 %) | 5 (16.7 %) | 21 (70 %) | 3 (10 %) | 1 (3.3 %) |
| Nevada | 1 (3.3 %) | 6 (20 %) | 19 (63.3 %) | 4 (13.3 %) | 0 (0 %) |
| New Hampshire | 0 (0 %) | 3 (10 %) | 23 (76.7 %) | 4 (13.3 %) | 0 (0 %) |
| New Jersey | 0 (0 %) | 9 (30 %) | 13 (43.3 %) | 6 (20 %) | 2 (6.7 %) |
| New Mexico | 2 (6.7 %) | 6 (20 %) | 16 (53.3 %) | 3 (10 %) | 3 (10 %) |
| New York | 0 (0 %) | 8 (26.7 %) | 15 (50 %) | 6 (20 %) | 1 (3.3 %) |
| North Carolina | 3 (10 %) | 4 (13.3 %) | 16 (53.3 %) | 5 (16.7 %) | 2 (6.7 %) |
| North Dakota | 1 (3.3 %) | 1 (3.3 %) | 24 (80 %) | 3 (10 %) | 1 (3.3 %) |
| Ohio | 1 (3.3 %) | 5 (16.7 %) | 18 (60 %) | 5 (16.7 %) | 1 (3.3 %) |
| Oklahoma | 2 (6.7 %) | 7 (23.3 %) | 10 (33.3 %) | 8 (26.7 %) | 3 (10 %) |
| Oregon | 0 (0 %) | 8 (26.7 %) | 16 (53.3 %) | 6 (20 %) | 0 (0 %) |
| Pennsylvania | 1 (3.3 %) | 7 (23.3 %) | 13 (43.3 %) | 7 (23.3 %) | 2 (6.7 %) |
| Puerto Rico | 0 (0 %) | 9 (30 %) | 16 (53.3 %) | 5 (16.7 %) | 0 (0 %) |
| Rhode Island | 0 (0 %) | 2 (6.7 %) | 26 (86.7 %) | 1 (3.3 %) | 1 (3.3 %) |
| South Carolina | 2 (6.7 %) | 7 (23.3 %) | 12 (40 %) | 7 (23.3 %) | 2 (6.7 %) |
| South Dakota | 4 (13.3 %) | 1 (3.3 %) | 19 (63.3 %) | 3 (10 %) | 3 (10 %) |

| **Jurisdiction** | **Large decrease**<br>**N (%)** | **Decrease**<br>**N (%)** | **Stable**<br>**N (%)** | **Increase**<br>**N (%)** | **Large increase**<br>**N (%)** |
|---|---|---|---|---|---|
| Tennessee | 0 (0 %) | 10 (33.3 %) | 14 (46.7 %) | 5 (16.7 %) | 1 (3.3 %) |
| Texas | 0 (0 %) | 8 (26.7 %) | 17 (56.7 %) | 5 (16.7 %) | 0 (0 %) |
| Utah | 0 (0 %) | 4 (13.3 %) | 24 (80 %) | 2 (6.7 %) | 0 (0 %) |
| Vermont | 0 (0 %) | 0 (0 %) | 30 (100 %) | 0 (0 %) | 0 (0 %) |
| Virginia | 1 (3.3 %) | 8 (26.7 %) | 15 (50 %) | 4 (13.3 %) | 2 (6.7 %) |
| Washington | 0 (0 %) | 4 (13.3 %) | 20 (66.7 %) | 6 (20 %) | 0 (0 %) |
| West Virginia | 4 (13.3 %) | 2 (6.7 %) | 18 (60 %) | 3 (10 %) | 3 (10 %) |
| Wisconsin | 0 (0 %) | 6 (20 %) | 15 (50 %) | 8 (26.7 %) | 1 (3.3 %) |
| Wyoming | 5 (16.7 %) | 0 (0 %) | 19 (63.3 %) | 0 (0 %) | 6 (20 %) |
